# Smartwatch-derived digital biomarkers distinguish episodic pain phenotypes in chronic low back pain

**DOI:** 10.64898/2026.09.02.26362006

**Authors:** Karim Khattab, Aniket Pratapneni, Gregorij Kurillo, Trisha Hue, Patricia Zheng, Jeffrey Lotz, Abel Torres-Espin, Jeannie F Bailey, The REACH Investigators

## Abstract

Chronic low back pain affects over half a billion people globally and is the leading cause of disability worldwide, yet objective tools for characterizing how individual patients experience pain over time remain lacking. Patients with chronic low back pain exhibit distinct pain trajectory patterns: some experience relatively stable pain while others experience episodic fluctuations with flare-ups. These trajectories have been found to be clinically meaningful but are currently captured only through subjective self-report. Consumer smartwatches offer an opportunity for passive, continuous, objective monitoring of physiological and behavioral signals that may reflect these fluctuations.

We evaluated whether temporal features extracted from six months of smartwatch-derived resting heart rate, heart rate variability, and step count data could discriminate between episodic and non-episodic pain phenotypes in 261 chronic low back pain patients from a longitudinal observational cohort. We applied stability-selected elastic-net logistic regression models to temporal features and evaluated model performance using nested cross-validation and SHAP-based feature importance analysis.

Summary statistics showed no significant differences between pain groups across any signal modality. Functional principal component analysis revealed high temporal heterogeneity in trajectories of resting heart rate, heart rate variability, and step count. Temporal trajectory models substantially outperformed summary-based approaches, with a models achieving areas under the receiver operating curve ranging from 0.66 to 0.82. Frequency-domain features and local variability measures drove classification performance, and combining heart rate and activity features provided complementary discriminative information, performing better than models trained on any subset of modalities.

These findings demonstrate the feasibility of passively collected smartwatch data as objective digital biomarkers for pain phenotyping in chronic low back pain, establishing a methodological approach that may permit personalized pain management through objective monitoring of patient pain and response to intervention.

**AUTHOR SUMMARY:** Chronic low back pain is the most common cause of disability worldwide, affecting over half a billion people. One of the biggest challenges in managing this condition is that patients experience it differently: some have relatively constant pain, while others experience unpredictable flare-ups. Understanding which pain trajectory a patient follows is important for guiding treatment but currently relies on patients recalling their experience, which is inherently subjective.

In this study, we asked whether data passively collected from consumer smartwatches could objectively distinguish between these two types of pain experience. We analyzed six months of heart rate and step count data from 261 chronic low back pain patients and found that simple averages of these signals looked identical between patient groups. However, when we examined how these signals changed and fluctuated over time, we were able to distinguish episodic from non-episodic pain patients with meaningful accuracy. Notably, combining heart rate and activity data performed better than either alone.

This work suggests that the smartwatch many people already wear on their wrist could one day help clinicians objectively monitor chronic pain in real time, reducing the burden of self-reporting and potentially enabling earlier detection of pain flare-ups.

## INTRODUCTION

Chronic low back pain (cLBP) is the most prevalent and costly cause of disability worldwide [1]. However, clinical phenotyping of cLBP patients remains challenging due to its multifactorial etiology and heterogeneous presentation [2]. While cLBP is often defined as “the persistence of pain beyond 3 months of symptoms” [3], the temporal dynamics of pain vary substantially between patients [4], [5], [6]. Some experience relatively stable pain levels over time, while others exhibit fluctuating, episodic patterns with distinct flare-ups [5], [7]. Patients often follow a specific pain trajectory over the time course of their low back pain, with little change in the nature of their trajectory [8]. Due to this stability and association with patient-specific measures, pain trajectories may represent more clinically meaningful phenotypes that could guide personalized treatment approaches [4].

Current methods for tracking pain trajectories rely heavily on patient recall and self-report, introducing subjectivity and recall bias [9]. The Visual Trajectories Questionnaire for Pain (VTQ-Pain) offers a validated, efficient approach to capturing self-reported pain trajectory type [10], but remains retrospective and subjective in nature. More broadly, patient-reported outcomes in cLBP such as pain intensity scales, disability indices, and quality of life measures, are administered episodically in clinical settings and cannot capture the dynamic, day-to-day fluctuations that characterize episodic pain. This represents a fundamental gap: the very feature that defines episodic pain (its temporal variability) is precisely what standard clinical assessment tools are least equipped to measure.

The emergence of wearable devices such as smartwatches presents an opportunity to develop objective, continuously monitored digital biomarkers for pain trajectory phenotyping. Digital health technologies are increasingly recognized for their potential to transform chronic pain management [11], [12]. Wearables enable passive collection of physiological and behavioral data in real-world settings, offering longitudinal resolution unattainable through traditional episodic clinical assessments [13], [14]. Consumer smartwatches in particular are already widely adopted, making them an attractive platform for scalable, low-burden monitoring.

Measures of resting heart rate (RHR) and heart rate variability (HRV) are associated with pain and have been shown to predict pain intensity [13], [14], [15]. Physical activity has been shown to exhibit complex, non-linear relationships with pain, particularly in cLBP [16]. However, these studies have focused on daily summary measures (such as daily averages of step count, sedentary time, or resting HRV) that do not capture temporal trends. Given that cLBP more often presents as an evolving condition than a static one, defined by temporal changes in intensity and pain flares, cross-sectional or aggregated measures are fundamentally mismatched to the phenomenon of interest. Additionally, day-to-day variability in activity has been associated with self-reported flare-ups in cLBP [17], and nighttime resting heart rate has been shown to predict next-day pain in chronic pain patients [18], suggesting that objective longitudinal measures may be more sensitive biomarkers of pain experience than static summaries. Critically, pain flares are complex, multidimensional events with physiological, autonomic, and behavioral manifestations that cannot be captured by any single signal domain. Combining temporally resolved heart rate and activity data may therefore provide a more comprehensive and robust proxy for episodic pain fluctuations than any individual data modality alone.

We hypothesized that objective temporal measures extracted from smartwatch-derived activity and heart rate trajectories are associated with pain trajectories characterized by episodic fluctuations. To test this, we first characterized the complexity of smartwatch trajectories across patients, anticipating that conventional summary measures would be insufficient for pain phenotype discrimination, motivating a richer, feature extraction-based approach. We then evaluated whether temporal trajectory features could discriminate between episodic and non-episodic pain phenotypes in cLBP patients using six months of smartwatch data from the BACKHOME UCSF cLBP cohort [19] (Fig. 1). We seek to demonstrate that temporal features from this data can reliably distinguish pain phenotypes, thus opening a path toward a form of scalable, objective monitoring of chronic pain in real-world settings that may enable earlier detection of flare-ups and support more personalized pain management.

**Figure 1:**
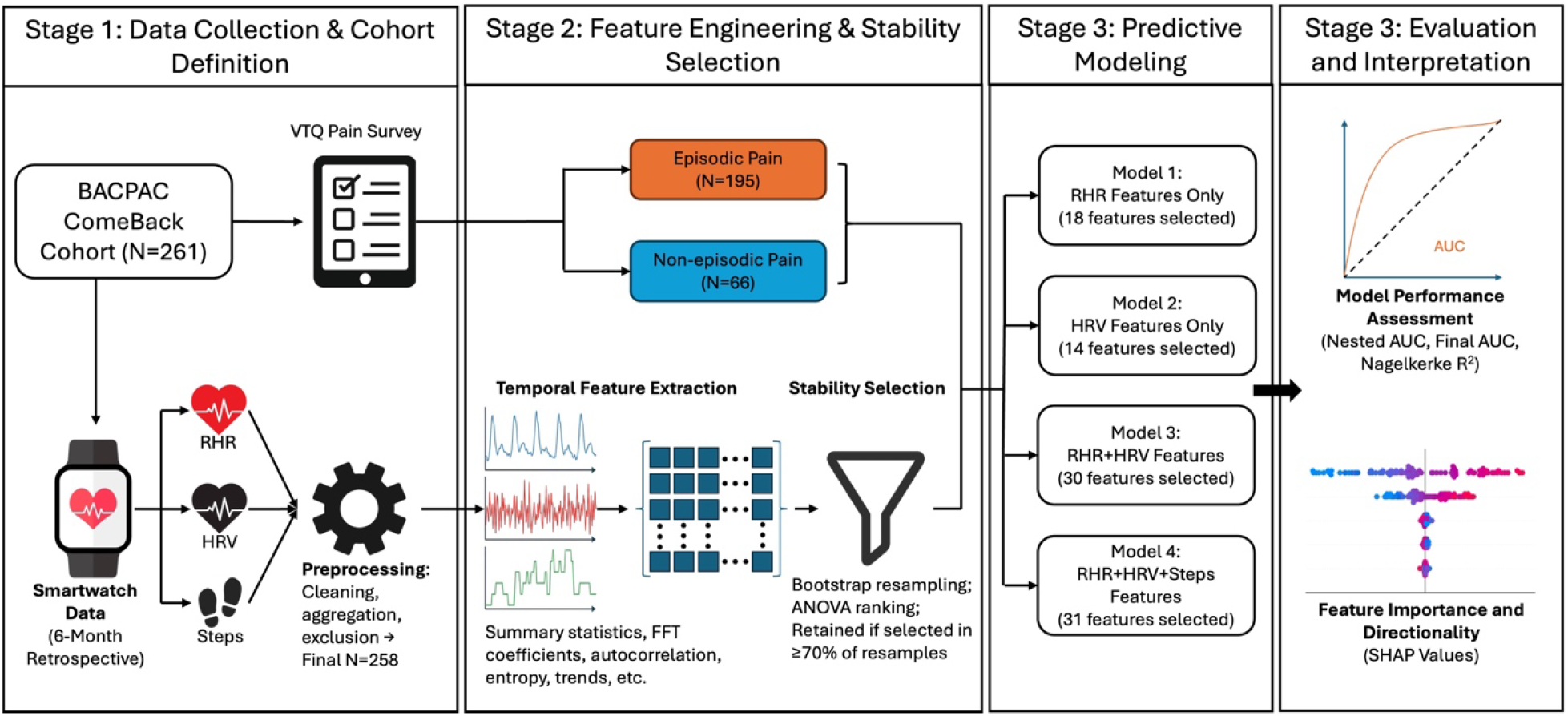
Study design and analytical pipeline.

**Figure 2:**
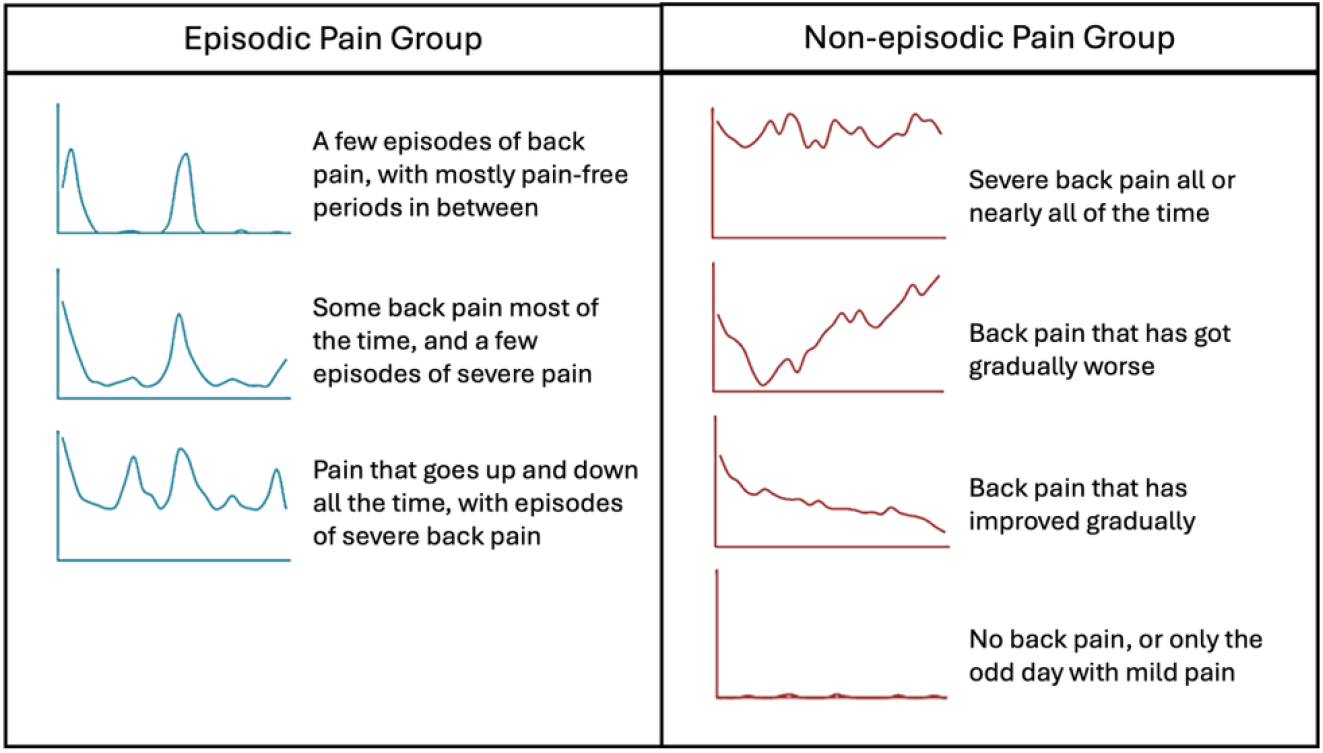
Episodic and non-episodic pain group classification from the Visual Trajectory Questionnaire for Pain. Patients were binned into two groups, episodic or non-episodic, based on their response to the VTQ-Pain questionnaire asking them to select the visual trajectory representing their pain in the past six months.

## RESULTS

### Cohort characteristics

261 cLBP patients (108 male, 153 female) from the BACKHOME [19] study cohort participated in the smartwatch study. Of those, 195 (74.7%) reported ‘episodic’ pain trajectories and 66 (25.3%) reported ‘non-episodic’ pain trajectories, with no significant group differences in age, sex distribution, or BMI. Across all subjects, the average resting heart rate was 64±8.3 bpm and the average heart rate variability was 29.1±10.4 ms. Of the 261 subjects, only 199 had daily step data, with an average daily step count of 5,472±2,653 steps.

### Trajectory complexity motivates feature-based classification

Conventional summary statistics including mean, standard deviation (SD), median, minimum, maximum, range, interquartile range, and coefficient of variation (CV) did not demonstrate a statistically significant difference between episodic and non-episodic groups across any modality (RHR, HRV, or Steps). PCA applied to 12 of these summary metrics (mean, SD, CV, and range for each of the three modalities) required 6 principal components to explain >90% of variance, indicating that even this compact summary representation lacks a simple, low-dimensional structure separating the groups. On average, cross-modality Pearson correlations were uniformly weak and had high relative error (RHR-HRV: r = –0.255±0.191, RHR-Steps: r = 0.109±0.173, HRV-Steps: r = –0.077±0.123), confirming that each modality captures largely independent physiological information.

Functional PCA (fPCA) of the full six-month trajectories revealed substantial temporal heterogeneity within each modality. To capture >90% of trajectory variance across patients, 6 functional principal components (fPCs) were required for RHR, 12 for HRV, and 34 for step count. The markedly higher complexity of step count trajectories reflects the greater day-to-day behavioral variability in physical activity relative to physiological signals. Together, these findings demonstrate that pain-relevant information is distributed across the temporal structure of wearable signals rather than concentrated in their average levels, motivating a broader temporal feature extraction approach (Fig. 3).

**Figure 3:**
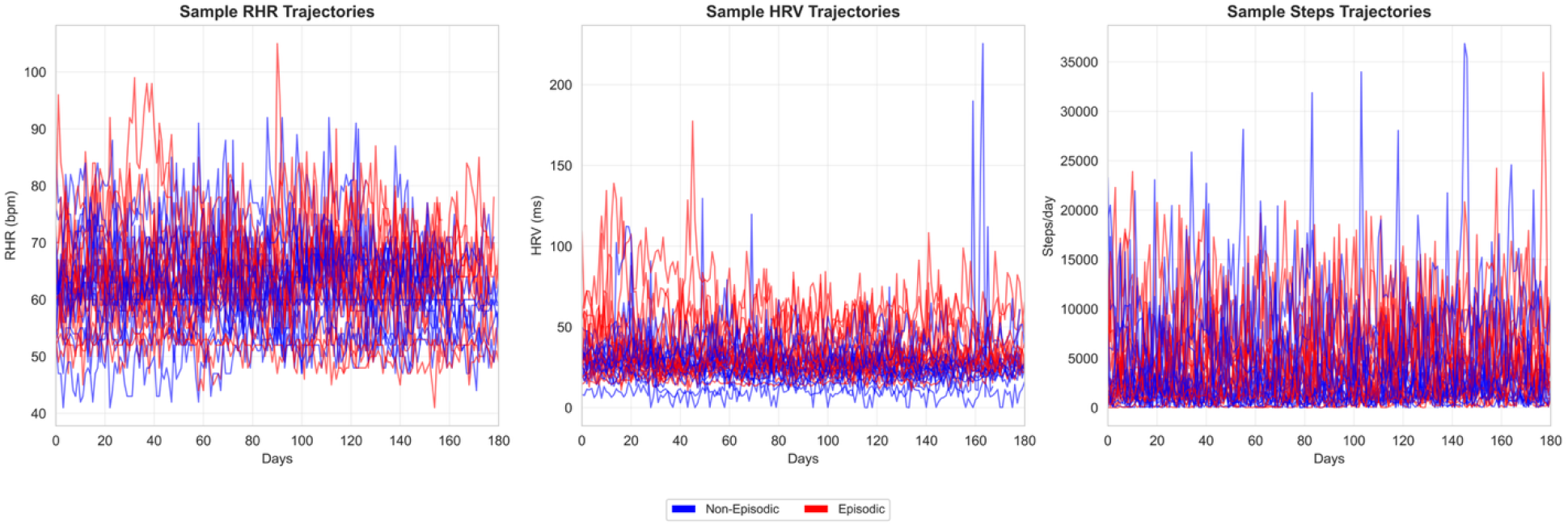
Sample trajectories illustrating signal complexity of RHR, HRV, and Step count data over the study period. 15 patients were selected at random from each of the Non-Episodic and Episodic cohorts to generate these plots.

### Discrimination of episodic vs. non-episodic pain phenotypes

To capture temporal aspects of the data with more granularity, we extracted 600 features from each data trajectory. After filtering these features for noncollinearity and stability, we applied four elastic-net regression models. These models were then evaluated using nested 3-fold cross validation (25 repeats) to predict episodic vs non-episodic pain classification. The RHR model (using only resting heart rate trajectory features) resulted in 18 selected features and achieved a nested AUC of 0.718±0.012 and final AUC of 0.718±0.012 (Nagelkerke R^2^=0.045). The HRV model (using only heart rate variability trajectory features) resulted in 14 selected features and performed more modestly, achieving a nested AUC of 0.668±0.024 and final AUC of 0.671±0.020 (Nagelkerke R^2^=-0.070). Combining RHR and HRV features (Heart model) resulted in 30 features and substantially improved performance, with a nested AUC of 0.781±0.019 and final AUC of 0.780±0.019 (Nagelkerke R^2^=0.28). The Heart + Steps model, incorporating features from all three variables, resulted in 31 features and achieved the best discrimination: nested AUC 0.816±0.029, final AUC 0.820±0.024 (Nagelkerke R^2^=0.48), demonstrating that activity trajectory features provide complementary information to heart rate metrics (Table 1).

**Table 1.** Model performance metrics.

| Model | Features (n) | Nested AUC (mean ± SD) | Final AUC (mean ± SD) | Nagelkerke $R^2$ |
| --- | --- | --- | --- | --- |
| RHR | 18 | $0.718 \pm 0.012$ | $0.718 \pm 0.012$ | 0.045 |
| HRV | 14 | $0.668 \pm 0.024$ | $0.671 \pm 0.020$ | -0.070 |
| Heart | 30 | $0.781 \pm 0.019$ | $0.780 \pm 0.019$ | 0.280 |
| Heart+Steps | 31 | $0.816 \pm 0.029$ | $0.820 \pm 0.024$ | 0.480 |

### Model feature importance

SHAP value analysis revealed that frequency-domain features (FFT coefficients) had the highest relative importance across all models. In the RHR model, the mean day-to-day change in RHR was also important, with lower day-to-day change associated with non-episodic pain (Fig. 4B). In the HRV model, the weekly autocorrelation and the weekly partial autocorrelation were important, with higher values in both associated with non-episodic pain (Fig. 4D). For both the RHR model and the HRV model, BMI was a relatively important feature with higher BMI values associated with non-episodic pain. In the Heart model, features with high relative importance included both RHR and HRV FFT features (Fig. 4F). The Heart + Steps model had RHR, HRV, and Steps features amongst the features with the highest relative importance (Fig. 4H).

**Figure 4:**
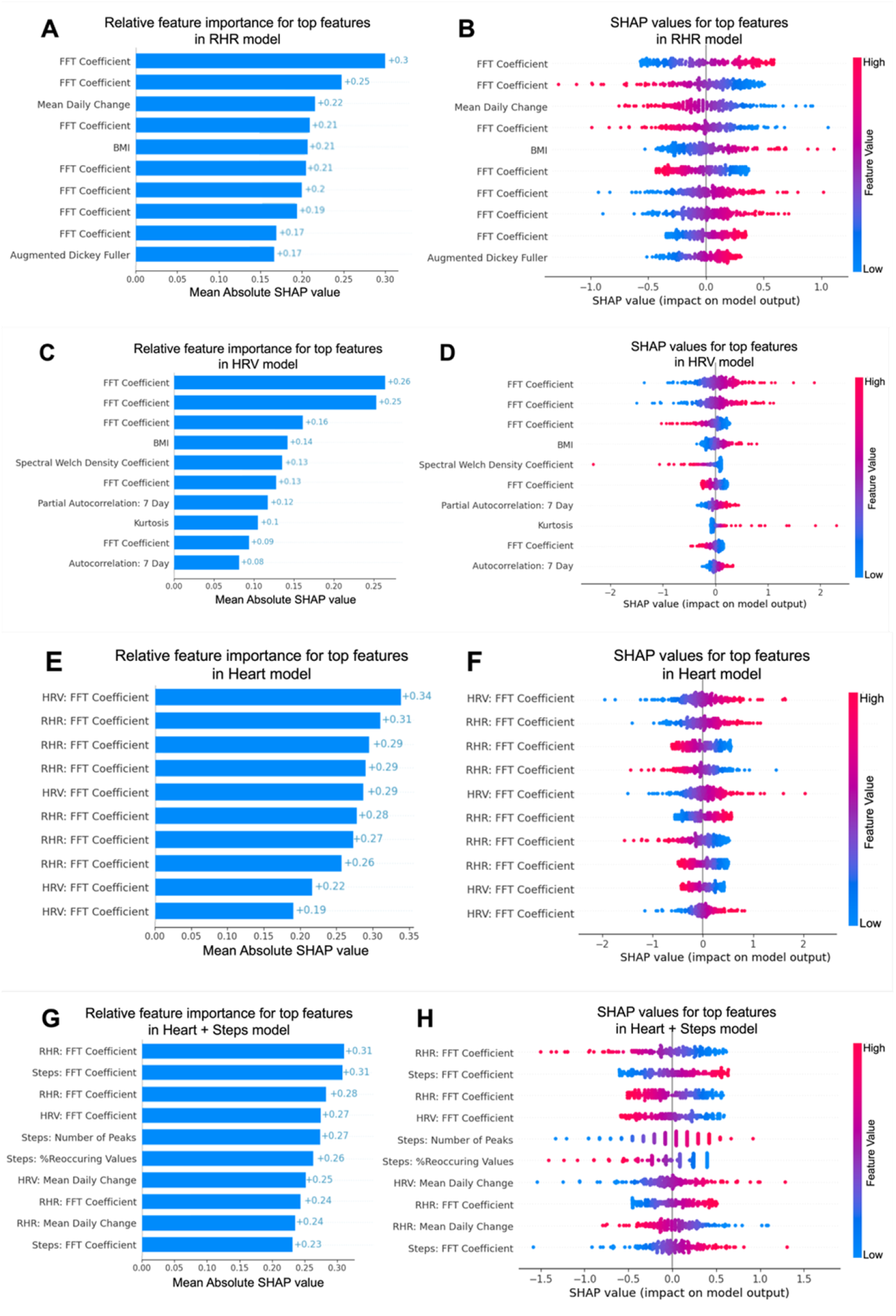
Relative feature importances and directions for the top ten features in the RHR (A, B), HRV (C, D), Heart (E, F), and Heart+Steps (G, H) models. A, C, E, G: Mean absolute SHAP values for the top 10 features in the RHR, HRV, Heart, and Heart+Steps models, respectively. B, D, F, H: SHAP values of the top ten features for each patient, color coded by feature value. Positive SHAP values represent relative importance for non-episodic pain group classification and negative SHAP values represent relative importance for episodic pain group classification.

## DISCUSSION

We demonstrate that temporal features extracted from consumer smartwatch data can discriminate between episodic and non-episodic pain phenotypes in cLBP patients. Crucially, this discriminative signal is not detectable using conventional summary statistics: mean, standard deviation, and related summary measures did not differ between pain groups across any modality (RHR, HRV, step count). It is only when the temporal structure of the trajectories is characterized through features capturing frequency content, day-to-day variability, and autocorrelation that meaningful group differences emerge. This finding has important methodological implications for future wearable-based pain research, suggesting that trajectory-level analyses may be necessary to detect associations that summary-based approaches miss.

The high temporal complexity revealed by fPCA further contextualizes this data complexity. Requiring 6, 12, and 34 functional principal components to explain 90% of variance in RHR, HRV, and step count trajectories, respectively, indicates that individual patients follow highly idiosyncratic physiological and behavioral patterns over time. This heterogeneity is particularly pronounced in step count, consistent with the substantial variability in how chronic pain patients navigate daily activity. The weak cross-modality correlations (all |r| < 0.26) confirm that heart rate and activity signals are largely independent, capturing different aspects of the patient experience, which likely explains why combining them improves model performance.

The combination of RHR, HRV, and activity trajectory features in the Heart + Steps model achieved the best discrimination (AUC = 0.820, R^2^ = 0.48). While overall model performance for the RHR- and HRV-only models was modest, combining these with step count trajectory features substantially improved model performance, suggesting that heart rate and activity trajectory features may explain different variances in pain classification. Heart rate metrics may reflect autonomic responses and stress-related processes [14], [15], while activity patterns may reflect functional capacity and pain-related movement avoidance[25]. While studies have shown that heart rate and activity may be changed in chronic pain patients [25], [26], [27], these signals are also sensitive to other stressors. Studies have shown that self-reported flare-ups in cLBP do not always coincide with increased pain intensity [28], [29], suggesting that other factors may influence this reporting. The multimodal approach may therefore provide robustness against such confounding factors, offering a more comprehensive picture of a patient’s chronic pain trajectory.

The prominence of frequency-domain and local variability features (FFT coefficients, day-to-day RHR change, weekly HRV autocorrelations) over global summary measures in SHAP analyses aligns with the fPCA findings and supports a coherent mechanistic interpretation: fluctuations at shorter timescales may better reflect the episodic nature of pain experience. Non-episodic pain patients exhibited lower day-to-day variability in RHR and higher weekly regularity in HRV, potentially suggesting a more stable autonomic state, while episodic patients showed more irregular physiological patterns, potentially reflecting the fluctuating physiological demands of intermittent pain episodes. These results corroborate prior findings that day-to-day variability in activity associates with flare-up incidence in cLBP and that nighttime heart rate predicts next-day pain in chronic pain conditions, extending these observations to the level of trajectory phenotyping [17], [18].

These findings have implications for digital health approaches to chronic pain management. Passively collected trajectory features could complement or partially substitute self-reported outcomes, reducing recall bias and patient burden. Real-time monitoring could enable earlier detection of impending flare-ups, facilitating timely clinical intervention. The use of consumer smartwatches enhances scalability and real-world applicability. However, this study is not without limitations. First, self-reported pain trajectories using the VTQ-Pain, while validated [10], remain susceptible to recall bias and do not allow for identification of specific flare-up incidences, instead relying on the assumption that patients reporting an episodic trajectory type are more likely to experience a higher frequency of flare-ups. Second, the class imbalance in our cohort (74.7% episodic) reflects the underlying clinical distribution but may limit model sensitivity for non-episodic classification; stratified sampling during feature selection and cross-validation was employed to mitigate this as much as possible. Third, while subjects with large portions of missing data were excluded, missing data persisted in the final dataset, as is inherent to passive wearable data collection, and this may have attenuated our ability to identify associations between features. Fourth, the retrospective nature of wearable data collection prevents causal inference about the relationship between trajectory features and pain phenotype. In addition, decreases in model performance may be related to the subjectivity and potential for bias in the VTQ-Pain rather than a lack of prediction power in the activity features. This further emphasizes the importance of objective measures for patient pain tracking and classification.

Future work will incorporate daily pain assessments, which would allow validation against specific flare-up events and enable trajectory features to be used for real-time prediction rather than retrospective classification. Integration of additional wearable-derived metrics (sleep quality, gait parameters, other activity metrics) may further improve phenotyping accuracy.

Extension of these methods to other chronic pain conditions would clarify whether observed patterns are specific to cLBP or generalizable across pain syndromes.

In conclusion, we demonstrate that temporal features extracted from RHR, HRV, and step count trajectories discriminate episodic from non-episodic cLBP pain phenotypes. This suggests that heart rate and activity tracking, especially in combination, may provide an opportunity for objective monitoring of patient pain, fluctuations, and response to interventions. This study also illustrates the temporal complexity of smartwatch data trajectories, and the failure of summary statistics to capture pain-relevant variation, highlighting the importance of trajectory-level analysis in wearable-based pain research. This work establishes a methodological foundation for the development of reliable, objective, and continuous digital biomarkers of patient pain experience.

## MATERIALS AND METHODS

### Ethics approval and informed consent

This study was approved by the University of California, San Francisco Institutional Review Board (institutional IRB #21-34420) and WCG IRB (IRB #20212455) as part of the multicenter BACPAC BACKHOME study. All participants provided written informed consent prior to enrollment. All study procedures were conducted in accordance with the Declaration of Helsinki and Good Clinical Practice guidelines. Participants consented to the retrospective collection and analysis of their Apple Watch health data from iPhone Healthkit.

### Study population

The study population was a subset of the BACKHOME [19] study cohort (, in which the key eligibility requirement was current chronic low back pain defined as pain between the lower posterior margin of the rib cage and the horizontal gluteal fold, which has persisted for at least the past 3 months and has resulted in pain on at least 50% of days in the past 6 months (per the NIH Task Force on Research Standards for cLBP [20], [21]), and that their low back pain was worse than any other bodily pain they were experiencing. BACKHOME participants who allowed access to their Apple Healthkit data and had enough data for analysis (≥120 of the 180 days available) were included.

### Data collection

With IRB approval and informed consent, cLBP patients in the BACPAC BACKHOME cohort completed the Visual Trajectories Questionnaire-Pain (VTQ-Pain) [10], a single question prompting patients to select the visual pain trajectory which best describes the course of their pain over the past 6 months (the only exclusion criterion was the inability to read or write English [19]). Patients were binned into ‘episodic’ or ‘non-episodic’ pain groups based on their response to the VTQ-Pain (Fig. 2). Then, for 261 participants who consented to iPhone Healthkit data access, we retrospectively collected resting heart rate, heart rate variability, and step count data, from iPhone HealthKit, for the six-month period described in the VTQ-Pain survey. We cleaned and aggregated the raw HealthKit data into daily average resting heart rate, daily median heart rate variability, and daily average step counts. From these daily values, we created six-month trajectories of RHR, HRV, and step count for each patient. Patients were excluded if they had more than 60 days of missing data in the 180-day time frame, resulting in a final cohort of 258 patients.

### Trajectory characterization

To evaluate the adequacy of summary statistics for group discrimination, we computed eight conventional summary metrics (mean, standard deviation, median, minimum, maximum, range, IQR, and coefficient of variation) for each modality and compared them between episodic and non-episodic groups using independent t-tests or Mann-Whitney U tests as appropriate. We further applied PCA to the 12-dimensional space of mean, SD, CV, and range across the three modalities. Cross-modality associations were assessed using Pearson correlation between modalities for each patient and then comparing the distributions of Pearson correlations in each group. To characterize the temporal complexity of each modality’s trajectory space, we applied functional PCA (fPCA) using the scikit-fda package, representing each patient’s trajectory as a continuous function prior to decomposition.

Group comparisons used independent t-tests for normally distributed continuous variables, Mann-Whitney U tests for non-normally distributed variables, and chi-square tests for categorical variables. All analyses were conducted in Python using numpy, pandas, scipy, scikit-learn, shap, and tsfresh libraries.

### Feature engineering

We applied the tsfresh package [22] to extract 600 time-series features per modality. Features included: summary statistics (mean, standard deviation, min/max, percentiles), distribution properties (skewness, kurtosis), linear trends, autocorrelation functions, spectral properties (FFT coefficients, entropy), complexity measures, and change quantifications. Features with missing values or zero variance across participants were removed. To address multicollinearity, we identified highly correlated feature pairs (>90%) and randomly removed one feature from each pair. This yielded a total of 300 features for model development.

We employed stability selection [23] to identify robust features for each model. Using 100 bootstrap resamples (50% sample size, stratified by outcome to maintain class imbalance), we ranked features by univariate ANOVA F-statistic and selected the statistically significant features (p<0.05) in each resample. Features selected in ≥70% of resamples were retained for final modeling. This yielded 18 RHR features, 14 HRV features, 30 Heart (RHR+HRV) features, and 31 Heart + Steps features.

### Statistical modeling

We trained elastic-net logistic regression models (scikit-learn) to predict episodic vs non-episodic classification using four feature sets: (1) RHR only, (2) HRV only, (3) Heart (RHR+HRV), and (4) Heart + Steps. Class imbalance was addressed through stratified sampling in all cross-validation folds.

Models were trained on a 70-30 training-testing split with a nested cross-validation. Both inner and outer cross-validations were 3-fold with 25 repeats. Hyperparameters were tuned in the inner fold using grid search with the final model hyperparameters taken from the best model across folds and repeats. For each model, we calculated the mean area under the receiver-operating curve (AUC) of the nested cross-validation (nested AUC), as well as the AUC of the final model refit following nested cross-validation. The final model coefficients and AUC were calculated from a single pass through the inner cross-validation using the full dataset. We report mean AUC across all outer folds (nested AUC) and final model AUC from refitting the full dataset with optimal hyperparameters.

Model performance was further assessed using Nagelkerke pseudo-R^2^, a scaled version of Cox-Snell R^2^ normalized to a maximum of 1, calculated from the final refitted model.

We computed SHAP (SHapley Additive exPlanations) values [24] using the shap package to quantify feature importance and direction of effect. SHAP values represent each feature’s contribution to individual predictions. We report mean absolute SHAP values (global feature importance) and SHAP value distributions stratified by feature values to visualize feature-outcome relationships.

## Data Availability

No data was generated by this study. Existing data was used from the UCSF BACKHOME cohort under the NIH BACPAC Research Program, available at https://doi.org/10.25934/PR00010819.

## ACKNOWLEDGEMENTS

The Back Pain Consortium (BACPAC) Research Program is administered by the National Institute of Arthritis and Musculoskeletal and Skin Diseases (NIAMS).

REACH Investigators: Research reported in this publication was supported by the National Institute of Arthritis and Musculoskeletal and Skin Diseases of the National Institutes of Health under Award Number U19AR076737. The content is solely the responsibility of the authors and does not necessarily represent the official views of the National Institutes of Health. The Core Center of Patient-centric, Mechanistic Phenotyping in Chronic Low Back (REACH) investigators include the following University of California, San Francisco (unless noted otherwise) personnel in alphabetical order: Zehra Akkaya, PhD; Prakruthi Amarkumar, PhD; Jeannie Bailey, PhD; Julia Barylak; Sigurd Berven, MD; Andrew Bishara, MD; Dennis M. Black, PhD; Noah Bonnheim, PhD; Atul Butte, MD, PhD; Jennifer Cummings; Karina Del Rosario, MD; Emilia Demarchis, MD; Sibel Demir-Deviren, MD; Susan K. Ewing, MS; Adam Ferguson, PhD; Aaron Fields, PhD; Scott M. Fishman, MD (University of California, Davis); Sergio Garcia Guerra; Fatemeh Gholi Zadeh Kharrat, PhD; Xiaojie (Summer) Guo; Misung Han, PhD; Trisha Hue, PhD; J. Russell Huie, PhD; C. Anthony Hunt, PhD; Anastasia Keller, PhD; Karim Khattab; Roland Krug, PhD; Gregorji Kurillo, PhD; Feng Lin; Thomas Link, MD, PhD; Jeffrey Lotz, PhD; John Lynch, PhD; Tong Lyu; Rob Matthew, PhD; Wolf Mehling, MD; Esmeralda Mendoza, MPH; Praveen Mummaneni, MD, MBA; Caroline Navy; Conor O’Neill, MD; Jessica Ornowski; Thomas Peterson, PhD; Ananya Rupanagunta (University of California, Berkeley); Aaron Scheffler, PhD, MS; Shalini Shah, MD (University of California, Irvine); Irina Strigo, PhD; Naoki Takegami, MD; Abel Torres-Espin, PhD (University of Waterloo); Salvatore Torrisi, PhD; Sachin Umrao, PhD; Rohit Vashisht, PhD; Joanna Veres; An (Joseph) Vu, PhD; Mark Steven Wallace, MD (University of California, San Diego); Lucy Ann Wu, MPH; Po-Hung Wu, PhD; Patricia Zheng, MD; Jiamin Zhou, MS.

The REACH Investigators would like to express their gratitude to the members of the UCSF clinical site research team, especially the Clinical Research Coordinators (CRCs) for their commitment and contributions toward the successful conduct of the study (in alphabetical order): Jamie Ahn^3^, Kristina Benirschke^1^, Alexandra Bryson^1^, Katherine Bunda^4^, Briana Davis^1^, Carolina Dorofeyev^2^, Rosalee Espiritu^4^, Pirooz Fereydouni^1^, Aamna Haq^1^, Nicholas Harris^1^, Sara Honardoost^3^, Gabriel Johnson^1^, Jennifer Johnson^1^, Edward Lingayo, Jr^2^, Robert Miller^3^, Phirum Nguyen^4^, Christopher Orozco^1^, Lindsay Ruiz-Graham^2^, Kie Shidara^1^, Kaitlyn Smith^1^, John (Boyuan) Xiao^1^, Michelle Yang^1^.

CRC Affiliations: ^1^University of California, San Francisco; ^2^University of California, Davis; ^3^University of California, Irvine; ^4^University of California, San Diego.

## DATA AVAILABILITY

The data that support the findings of this study are not openly available due to reasons of sensitivity and are available from the corresponding author upon reasonable request. Data are located in controlled access data storage at the University of California, San Francisco.

